# Glycaemic Burden and Cognitive Screen-Positive Status in Younger Indian Adults with Long-Standing Type 2 Diabetes: A Cross-Sectional Study with a MoCA Cutoff Sensitivity Analysis in a Lower-Education Population

**DOI:** 10.64898/2026.09.22.26363620

**Authors:** Siddharth Manish Lodha, Sumit Kumar, Shubho Acharya, Ritvik Baweja, Sandeep Garg

**Author notes:** Corresponding author: Dr. Siddharth Manish Lodha, Department of Internal Medicine, Maulana Azad Medical College, Bahadur Shah Zafar Marg, New Delhi, 110002, India. Joint first authors- Dr. Siddharth Manish Lodha and Dr. Sumit Kumar. email id.

## Abstract

**Background:** Structured cognitive screening in diabetes is recommended only from age 65, leaving younger adults with prolonged type 2 diabetes mellitus (T2DM) exposure unmonitored. We quantified the association of HbA1c and diabetes duration with Montreal Cognitive Assessment (MoCA) screen-positive status in adults aged ≤ 60 years with T2DM of ≥ 5 years, and examined against matched controls how the MoCA cutoff (< 26, < 24, < 23) affects the group contrast in a lower-education population.

**Methods:** Cross-sectional study at a tertiary-care centre in North India: 110 adults with T2DM and 50 controls frequency-matched for age, sex, and education. Within diabetics, HbA1c and duration were modelled continuously by modified Poisson regression at each pre-specified cutoff and by Spearman correlation with MoCA total score. Prevalence ratios (PR) for diabetics versus controls at each cutoff were adjusted for age, sex, education, BMI, and hypertension.

**Results:** Within diabetics, MoCA total score correlated with HbA1c (ρ = −0.456) and duration (ρ = −0.318; both p < 0.001). Screen-positive prevalence rose 5% per 1% HbA1c at < 26 (PR 1.05, 95% CI 1.02-1.09), 12% at < 24 (PR 1.12, 1.06-1.18), and 17% at < 23 (PR 1.17, 1.09-1.26), and 11% per year of duration at < 23 (PR 1.11, 1.06-1.17). Median education was 10 and 9 years; 76.0% of controls screened positive at < 26, and the diabetes association was cutoff-dependent: adjusted PR 1.08 (0.89-1.29) at < 26, 1.70 (1.10-2.63) at < 24, and 6.72 (2.23-20.23) at < 23, where 44.5% of diabetics versus 6.0% of controls screened positive (absolute difference 38.5 percentage points, 95% CI 25.0 to 48.7).

**Conclusions:** In working-age adults with long-standing T2DM, screen-positive prevalence rose continuously with HbA1c at every cutoff, a glycaemic signal independent of threshold choice. The standard < 26 cutoff flagged over three-quarters of both groups and could not triage; < 23 separated diabetics from controls. Screen-positive status is not a diagnosis; MoCA triage at a population-appropriate cutoff in this group warrants prospective validation against a neuropsychological reference standard.

## INTRODUCTION

Type 2 diabetes mellitus (T2DM) affects approximately 589 million adults worldwide, projected to reach 853 million by 2050 [1]; India harbours the second-largest diabetic population, with a prevalence of 11.4% [2]. T2DM is an established risk factor for cognitive dysfunction: pooled relative risks for dementia subtypes range from 1.56 to 2.27 [3], the summary RR for dementia is 1.59 (95% CI 1.40-1.80) [4], and the largest domain-specific deficits are in processing speed (d = −0.33), executive function (d = −0.33), and verbal memory (d = −0.28) [5]. Proposed mechanisms include cerebral microvascular injury, brain insulin resistance, and hyperglycaemia-driven neuroinflammation [6–9]. Earlier onset carries higher dementia risk (HR 1.24 per 5-year earlier onset) [17], and because mild cognitive impairment is a transitional state during which intervention may slow progression to dementia [8,10], early identification of adults who accumulate long diabetes exposure before old age is a clinical priority.

Current guidance leaves this group unmonitored. The ADA recommends monitoring cognitive capacity across the life span, but structured screening for mild cognitive impairment or dementia only from age 65, at the initial visit and annually thereafter [13]. Consistent with this, studies of diabetes-associated cognitive impairment have predominantly enrolled elderly populations (≥ 60 or ≥ 65 years), including the RECOGNISED consortium [11], the ACCORD-MIND trial [12], and recent meta-analyses [34–35]. Indian studies have examined cognitive impairment in T2DM across age groups [14–16], but none required a minimum diabetes duration, so the contribution of glycaemic burden and long-standing disease (≥ 5 years) in working-age adults remains unquantified.

What screening can achieve in this group is limited by what the instrument is. The MoCA is a screening instrument, not a diagnostic tool [18]: a score below a cutoff establishes screen-positive status, whereas a diagnosis of mild cognitive impairment or dementia requires comprehensive neuropsychological assessment together with a subjective cognitive complaint and evaluation of functional status [10]. Current diabetes guidance frames brief instruments in the same terms, reserving formal assessment for those who screen positive [13]. The purpose of the MoCA here is therefore triage, and a cutoff is useful only if it partitions a population into a group worth evaluating further and a group that is not. Judged against that standard, the conventional cutoff of < 26 proposed by Nasreddine et al. [18] fails in lower-education populations. In the US National Alzheimer’s Coordinating Center database (n = 3,895), optimal cutoffs for MCI were 25, 24, and 23 for non-Hispanic Whites, Hispanics, and non-Hispanic Blacks, respectively [19]; among African Americans with T2D (n = 415), 93.5% screened positive at < 26 [20]; in a community-based African American sample, 80% of neurologically healthy participants fell below this threshold [21]; and in a Swedish normative cohort (n = 758), 37.3% of healthy elderly adults did so [22]. In India, normative data showed a mean MoCA of 19.4, with < 26 misclassifying 78.2% as impaired [23], and the ICMR-NCTB validation found all language-specific cutoffs fell below < 26 [24]. Carson et al. recommended < 23 to maximise overall accuracy [25]. Any study of screen-positive status in such a population must therefore show that its findings do not depend on the threshold chosen.

Three gaps in the literature follow. First, exposure-response gradients for HbA1c and diabetes duration have not been quantified as per-unit prevalence ratios in younger adults with long-standing disease, nor tested for robustness across cutoffs; most cross-sectional diabetes-cognition studies apply a single cutoff without systematic comparison [26–27]. Second, studies in this field either lacked a non-diabetic control group [15–16,27], without which false-positive rates cannot be estimated, or relied on logistic regression [14], whose odds ratios overstate associations when outcome prevalence exceeds 20-30% [28]; modified Poisson regression, the recommended approach for common outcomes in cross-sectional studies [28–30], has not been applied. Third, the evidence base is drawn predominantly from elderly cohorts [11–12,33–35].

The primary objective was therefore to quantify the association of HbA1c and diabetes duration with screen-positive cognitive impairment and with MoCA score in adults aged ≤ 60 years with T2DM of ≥ 5 years, estimating per-unit prevalence ratios at three pre-specified cutoffs (< 26, < 24, < 23) to establish whether the gradients are robust to threshold choice. Secondary objectives were to quantify screen-positive prevalence in diabetics versus frequency-matched non-diabetic controls at each cutoff, and thereby to determine how cutoff choice affects the instrument’s capacity to separate the groups; to compare MoCA total and subdomain scores; and to examine associations of fasting lipid fractions with MoCA. The study was designed to characterise the glycaemic signal and to inform the choice of a triage threshold, not to validate one against a diagnostic reference standard; no gold-standard neuropsychological battery was administered.

## METHODS

### Study Design, Setting, and Participants

This hospital-based analytical cross-sectional study was conducted at a tertiary-care centre in North India between October 2025 and April 2026. Adults aged ≤ 60 years with T2DM of ≥ 5 years’ duration (n = 110) were recruited consecutively from the diabetes/internal medicine outpatient clinic; all eligible patients attending during the study period were invited. A non-diabetic comparison group (n = 50), frequency-matched for age, sex, and education, was recruited concurrently from adults accompanying patients to the hospital. Non-diabetic status was confirmed by HbA1c < 5.7% (39 mmol/mol) with no prior diabetes diagnosis or antidiabetic medication; individuals with prediabetes (HbA1c 5.7-6.4% [39-46 mmol/mol]) were excluded from the comparison group. Exclusion criteria for both groups were prior dementia or major neurocognitive disorder, stroke with residual aphasia, severe neurological or psychiatric illness, acute metabolic decompensation, hypothyroidism, history of alcohol abuse, severe anaemia, severe sensory impairment precluding testing, and medications known to substantially affect cognition. No formal a priori sample-size calculation was performed; the sample reflects all eligible participants enrolled in the study period, and all estimates, particularly the cutoff-specific prevalence ratios, are interpreted as hypothesis-generating with precision conveyed by the confidence intervals. The study is reported in accordance with the STROBE guidelines for cross-sectional studies. Clinical trial number: not applicable

### Data Collection and Cognitive Assessment

Data were collected using a structured proforma. Demographic and clinical variables included age, sex, years of education, BMI (calculated from measured height and self-reported weight), hypertension, and diet; diabetes duration and treatment regimen were additionally recorded for diabetics. HbA1c and fasting lipid profile were measured in both groups. The primary exposures were HbA1c (per 1% increment) and diabetes duration (per year). The MoCA was administered in a quiet room in the validated Hindi or English version, yielding a maximum score of 30; the standard +1 education correction was applied for ≤ 12 years of formal education [18]. The primary outcome was screen-positive status, defined at three pre-specified cutoffs: < 26 (original validation [18], retained for comparability with prior Indian studies [31]); < 24 (an intermediate threshold between the standard cutoff and the lower values recommended for non-Western and lower-education populations [25,32–33]); and < 23 (the optimal sensitivity-specificity trade-off identified by Carson et al. [25] and independently validated in Indian T2DM by Gupta et al. at 69.2% sensitivity and 71.8% specificity [33]). Secondary outcomes were MoCA total and subdomain scores.

### Statistical Analysis

Analyses were performed in R (version 4.6.0). Continuous variables were summarised as mean ± SD or median (IQR) and categorical variables as frequencies and percentages. Between-group comparisons used the Mann-Whitney U test (continuous) and the chi-square or Fisher’s exact test (categorical); non-parametric tests were selected because Shapiro-Wilk testing indicated significant departures from normality for all continuous variables (all p < 0.05). Equality of variances in MoCA total score between groups was assessed by the Levene test. For the primary analysis, within the diabetic group (n = 110), HbA1c and duration were modelled continuously by modified Poisson regression with robust standard errors [29], yielding the prevalence ratio (PR) for screen-positive status per 1% HbA1c and per year of duration at each cutoff; log-linearity was assessed by adding quadratic terms to the crude models (neither HbA1c², p = 0.077, nor duration², p = 0.149, reached significance, supporting the linear specification). Spearman’s ρ quantified threshold-free correlations of MoCA total and subdomain scores with HbA1c and duration. For the secondary group contrast, the PR for screen-positive status in diabetics versus controls was estimated at each cutoff by modified Poisson regression, the recommended approach for cross-sectional studies with common binary outcomes [28,30]; multivariable models adjusted for age, sex, education, BMI, and hypertension (n = 160), and a sensitivity analysis additionally adjusted for fasting lipid fractions (n = 136; lipid data were available for 86 of 110 diabetics and all 50 controls). Absolute differences in screen-positive prevalence (risk differences) were reported with 95% confidence intervals by Newcombe’s method. Associations of lipid fractions with MoCA total score were examined by multiple linear regression adjusted for age, sex, education, BMI, hypertension, and diabetes status (n = 136). Subdomain comparisons and lipid analyses were exploratory and not corrected for multiple testing. A two-tailed p < 0.05 was considered significant.

## RESULTS

### Participants

A total of 160 participants were enrolled: 110 with T2DM and 50 non-diabetic controls (Table 1). Groups were comparable in age (median 50 [46–55] vs. 50 [46–53] years), sex (50.0% vs. 52.0% male), and education (median 10 [7–12] vs. 9 [8-10.75] years). Diabetics had higher BMI (median 24 vs. 23 kg/m²; p = 0.001), more hypertension (39.1% vs. 8.0%; p < 0.001), higher LDL and triglycerides, and lower HDL (all p ≤ 0.038). Median HbA1c in diabetics was 8.5% (69 mmol/mol) [IQR 7.6-10.55], mean 9.25 ± 2.09% (78 ± 23 mmol/mol), and median diabetes duration 7 [6–10] years.

**Table 1.**
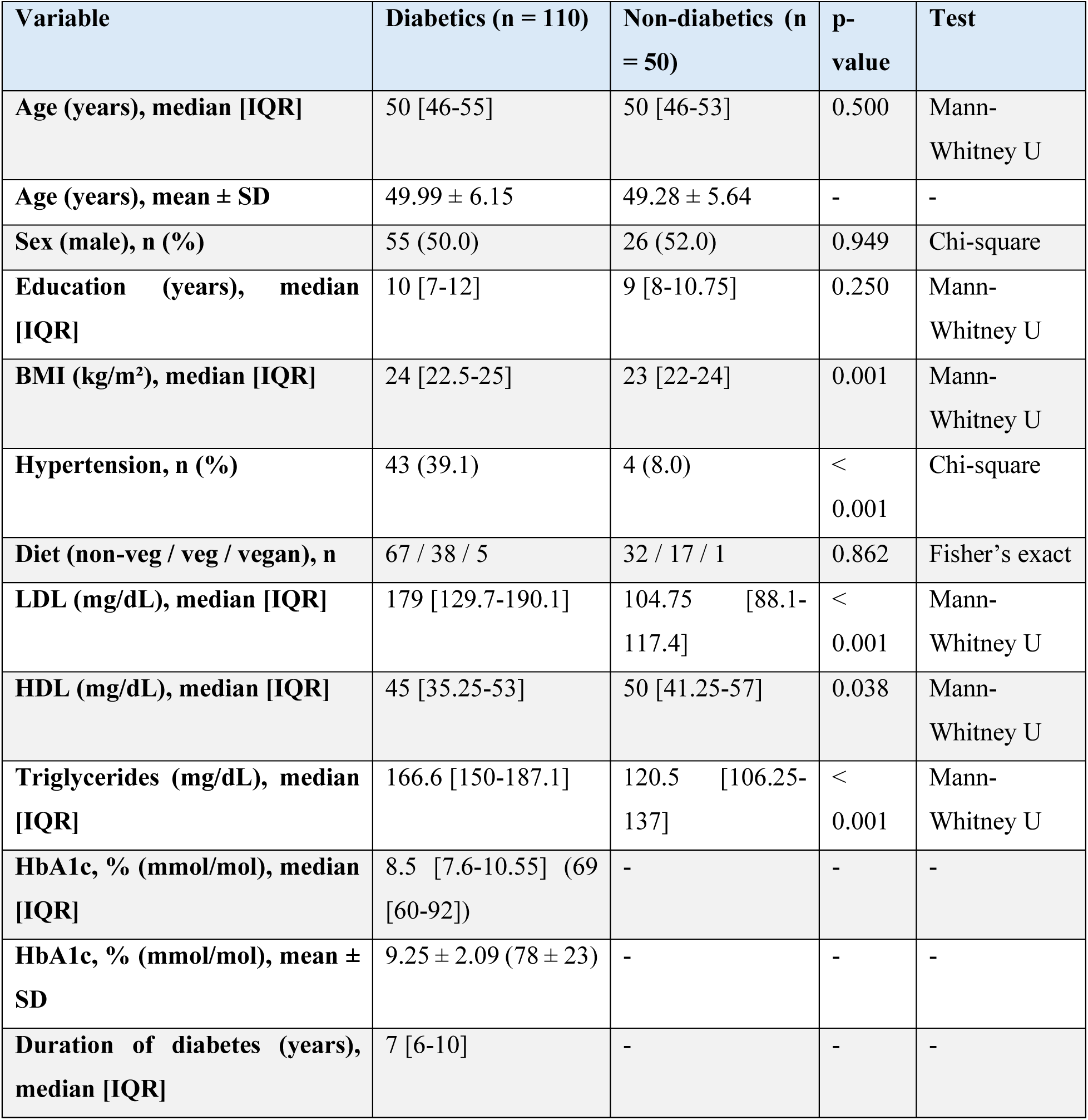
Sociodemographic and clinical characteristics of study participants (N = 160)

| Variable | Diabetics (n = 110) | Non-diabetics (n = 50) | p-value | Test |
| --- | --- | --- | --- | --- |
| Age (years), median [IQR] | 50 [46-55] | 50 [46-53] | 0.500 | Mann-Whitney U |
| Age (years), mean ± SD | 49.99 ± 6.15 | 49.28 ± 5.64 | - | - |
| Sex (male), n (%) | 55 (50.0) | 26 (52.0) | 0.949 | Chi-square |
| Education (years), median [IQR] | 10 [7-12] | 9 [8-10.75] | 0.250 | Mann-Whitney U |
| BMI (kg/m<sup>2</sup>), median [IQR] | 24 [22.5-25] | 23 [22-24] | 0.001 | Mann-Whitney U |
| Hypertension, n (%) | 43 (39.1) | 4 (8.0) | < 0.001 | Chi-square |
| Diet (non-veg / veg / vegan), n | 67 / 38 / 5 | 32 / 17 / 1 | 0.862 | Fisher's exact |
| LDL (mg/dL), median [IQR] | 179 [129.7-190.1] | 104.75 [88.1-117.4] | < 0.001 | Mann-Whitney U |
| HDL (mg/dL), median [IQR] | 45 [35.25-53] | 50 [41.25-57] | 0.038 | Mann-Whitney U |
| Triglycerides (mg/dL), median [IQR] | 166.6 [150-187.1] | 120.5 [106.25-137] | < 0.001 | Mann-Whitney U |
| HbA1c, % (mmol/mol), median [IQR] | 8.5 [7.6-10.55] (69 [60-92]) | - | - | - |
| HbA1c, % (mmol/mol), mean ± SD | 9.25 ± 2.09 (78 ± 23) | - | - | - |
| Duration of diabetes (years), median [IQR] | 7 [6-10] | - | - | - |

### Glycaemic Burden, Diabetes Duration, and Screen-Positive Status

Within diabetics, MoCA total score correlated negatively with HbA1c (ρ = −0.456) and with diabetes duration (ρ = −0.318; both p < 0.001; Figure 1). Each 1% increase in HbA1c was associated with 5% higher screen-positive prevalence at < 26 (PR 1.05, 1.02-1.09; p = 0.004), 12% at < 24 (PR 1.12, 1.06-1.18; p < 0.001), and 17% at < 23 (PR 1.17, 1.09-1.26; p < 0.001; Table 2). Each additional year of diabetes duration was not associated with screen-positive status at < 26 (PR 1.01, 0.98-1.04; p = 0.45) but was associated with 6% higher prevalence at < 24 (PR 1.06, 1.02-1.10; p = 0.005) and 11% at < 23 (PR 1.11, 1.06-1.17; p < 0.001). At the subdomain level, HbA1c correlated with abstraction (ρ = −0.354, p < 0.001), delayed recall (ρ = −0.308, p = 0.001), visuospatial/executive function (ρ = −0.260, p = 0.006), and attention (ρ = −0.234, p = 0.014) but not with naming, language, or orientation; duration correlated only with delayed recall (ρ = −0.247, p = 0.009), with a non-significant trend for attention (ρ = −0.182, p = 0.056).

**Fig. 1.**
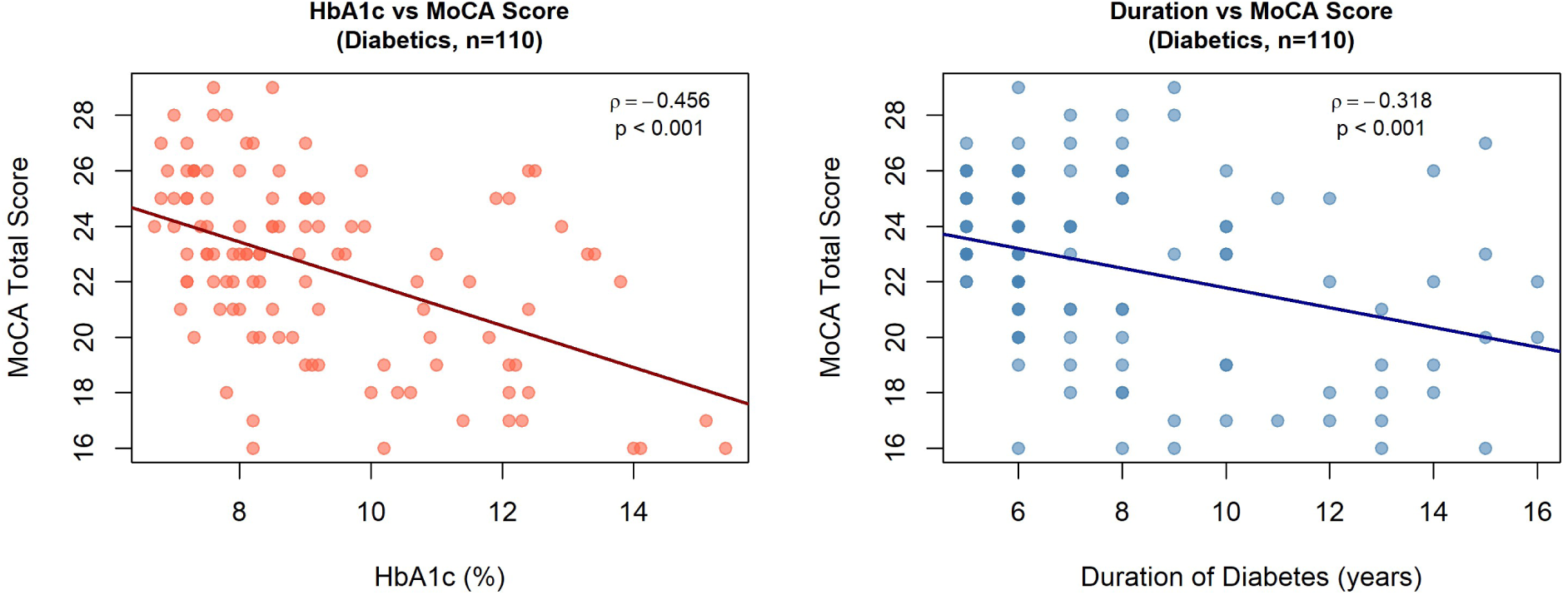
Scatter plots of MoCA total score against HbA1c and against diabetes duration, with Spearman correlation coefficients (diabetics, n = 110)

**Table 2.** Within-diabetic prevalence ratios of screen-positive cognitive impairment per 1-unit increase in HbA1c and per 1-year increase in diabetes duration across three MoCA cutoffs (modified Poisson regression with robust standard errors; diabetic subgroup, n = 110)

| Within-diabetic predictor, by MoCA cutoff | PR per unit | 95% CI | p-value |
| --- | --- | --- | --- |
| HbA1c (per 1% increase), < 26 | 1.05 | 1.02-1.09 | 0.004 |
| HbA1c (per 1% increase), < 24 | 1.12 | 1.06-1.18 | < 0.001 |
| HbA1c (per 1% increase), < 23 | 1.17 | 1.09-1.26 | < 0.001 |
| Diabetes duration (per 1 year), < 26 | 1.01 | 0.98-1.04 | 0.45 |
| Diabetes duration (per 1 year), < 24 | 1.06 | 1.02-1.10 | 0.005 |
| Diabetes duration (per 1 year), < 23 | 1.11 | 1.06-1.17 | < 0.001 |

### MoCA Score Distribution and Group Contrast by Cutoff

Mean MoCA was lower in diabetics than controls (22.49 ± 3.24 vs. 24.32 ± 1.50; p < 0.001), and the control mean itself lay below the standard cutoff of 26. Dispersion was greater in diabetics (variance 10.5 vs. 2.3; Levene test p < 0.001); the lowest score recorded in any control was 21, and 30 of 110 diabetics (27.3%) scored below this value. The between-group association was strongly cutoff-dependent (Table 3). At < 26, 80.9% of diabetics and 76.0% of controls screened positive, an absolute difference of 4.9 percentage points (95% CI −7.9 to +19.7; crude PR 1.06, 95% CI 0.89-1.27; p = 0.496). At < 24, prevalence was 59.1% versus 32.0% (difference 27.1 percentage points, 95% CI 10.4 to 41.3; PR 1.85, 1.20-2.85; p = 0.005). At < 23, prevalence was 44.5% versus 6.0% (difference 38.5 percentage points, 95% CI 25.0 to 48.7; Fisher’s exact p < 0.001; crude PR 7.42, 2.43-22.68). Adjustment for age, sex, education, BMI, and hypertension gave PRs of 1.08 (0.89-1.29) at < 26, 1.70 (1.10-2.63; p = 0.018) at < 24, and 6.72 (2.23-20.23; p < 0.001) at < 23. Additional adjustment for lipids (n = 136) left the < 26 and < 23 estimates essentially unchanged (aPR 1.05 and 6.81) and the < 24 estimate modestly higher (aPR 1.98). Because only 3 of 50 controls screened positive at < 23, the ratio at this cutoff is imprecise and is best read alongside the absolute difference, which does not depend on a three-event denominator.

**Table 3.** Crude and adjusted prevalence ratios for screen-positive cognitive impairment by MoCA cutoff (< 26, < 24, < 23), comparing T2DM with non-diabetic controls (modified Poisson regression with robust standard errors; base model adjusted for age, sex, education, BMI, hypertension; sensitivity model additionally adjusted for LDL, HDL, triglycerides)

| MoCA cutoff | Diabetics screening positive, n/N (%) | Non-diabetics screening positive, n/N (%) | Crude PR (95% CI) | Adjusted PR (95% CI)† | Adjusted PR + lipids (95% CI)‡ |
| --- | --- | --- | --- | --- | --- |
| < 26 | 89/110 (80.9) | 38/50 (76.0) | 1.06 (0.89-1.27) | 1.08 (0.89-1.29) | 1.05 (0.82-1.35) |
| < 24 | 65/110 (59.1) | 16/50 (32.0) | 1.85 (1.20-2.85)** | 1.70 (1.10-2.63)* | 1.98 (1.27-3.10)** |
| < 23 | 49/110 (44.5) | 3/50 (6.0) | 7.42 (2.43-22.68)*** | 6.72 (2.23-20.23)*** | 6.81 (2.43-19.12)*** |

† Adjusted for age, sex, education, BMI, hypertension (n = 160). ‡ Additionally adjusted for LDL, HDL, triglycerides (complete-case n = 136). \* p < 0.05; \*\* p < 0.01; \*\*\* p < 0.001. PR estimated by modified Poisson regression with robust standard errors [29].

### Exploratory Analyses

Diabetics scored lower in abstraction (1.08 ± 0.79 vs. 1.42 ± 0.61; p = 0.013) and delayed recall (2.45 ± 1.44 vs. 2.96 ± 0.83; p = 0.008), with no significant difference in the other five subdomains (Table 4). In adjusted linear models (n = 136), HDL was weakly positively associated with MoCA total score (β = +0.050; p = 0.039); LDL and triglycerides showed no association. Screen-positive prevalence at < 23 did not differ by sex (43.6% vs. 45.5%; p = 1.0).

**Table 4.** MoCA subdomain scores between diabetics and non-diabetics.

| **Subdomain (max score)** | **Diabetics, median [IQR]** | **Non-diabetics, median [IQR]** | **Diabetics, mean ± SD** | **Non-diabetics, mean ± SD** | **p-value** |
| --- | --- | --- | --- | --- | --- |
| **Visuospatial/executive (5)** | 4 [3-5] | 4 [3-5] | 3.66 ± 1.05 | 3.98 ± 0.77 | 0.097 |
| **Naming (3)** | 3 [3-3] | 3 [3-3] | 2.82 ± 0.43 | 2.84 ± 0.37 | 0.920 |
| **Attention (6)** | 4 [3-6] | 4 [4-5] | 3.90 ± 1.86 | 4.34 ± 0.75 | 0.439 |
| **Language (3)** | 2 [2-2] | 2 [2-2] | 1.96 ± 0.62 | 2.10 ± 0.51 | 0.209 |
| **Abstraction (2)** | 1 [0-2] | 1 [1-2] | 1.08 ± 0.79 | 1.42 ± 0.61 | 0.013 |
| **Delayed recall (5)** | 2 [2-3] | 3 [2-3.75] | 2.45 ± 1.44 | 2.96 ± 0.83 | 0.008 |
| **Orientation (6)** | 6 [6-6] | 6 [6-6] | 5.85 ± 0.39 | 5.78 ± 0.42 | 0.259 |

*All comparisons by Mann-Whitney U test (non-parametric, Shapiro-Wilk $p < 0.05$ for all subdomain distributions).*

## DISCUSSION

In working-age adults with long-standing T2DM in a lower-education population, screen-positive cognitive status rose continuously with glycaemic burden. The HbA1c gradient was present at every cutoff examined, from PR 1.05 per 1% at < 26 to 1.17 at < 23, and the threshold-free correlation between HbA1c and MoCA total score (ρ = −0.456) confirms that the signal does not depend on where a cutoff is drawn. Diabetes duration showed a parallel but weaker gradient that emerged only at the lower cutoffs. Against matched controls, the same population illustrated why cutoff choice matters: at the standard < 26 threshold, 76.0% of healthy controls and 80.9% of diabetics screened positive, a yield that would refer almost the entire clinic onward and separates neither group from the other, whereas at < 23, 44.5% of diabetics versus 6.0% of controls screened positive, an absolute excess of 38.5 percentage points (95% CI 25.0 to 48.7).

The glycaemic gradient is the study’s most robust finding because it is internal to the diabetic group and independent of both the comparison group and the threshold. It is consistent with the ACCORD-MIND cross-section (1% higher HbA1c, 1.75 points lower on Digit Symbol Substitution) [12,40], with a 2025 meta-analysis identifying HbA1c > 9% (75 mmol/mol) and duration of 8-9 years as critical thresholds [35], the former exceeded at our cohort mean of 9.25% (78 mmol/mol), and with the UK Biobank analysis in which T2DM-related executive deficits and grey-matter atrophy worsened with duration [42]. At the population level, the largest longitudinal meta-analysis (40 studies, > 7 million participants) found a clear duration gradient for incident dementia (HR about 1.4 for ≥ 5 years; 2.76 for ≥ 15 years) but a modest, non-significant continuous HbA1c association (HR 1.18, 95% CI 0.97-1.45) [41]; a cross-sectional screening gradient in a poorly controlled clinic population is not in conflict with that, but neither does it establish that glycaemic control causes the difference. Cross-sectional data cannot separate forward from reverse causation, and the ACCORD-MIND finding that intensive lowering to < 6% (42 mmol/mol) did not improve cognition [12] cautions against inferring that glycaemic intensification would reverse the association observed here. That the gradient appears at a mean age of 50 with median duration of 7 years, in the group that age-based screening guidance currently excludes [13], is the observation with clinical relevance.

The saturation of < 26 was anticipated and is not India-specific: it matches the 78.2% of cognitively normal Kerala adults classified as impaired by Iype et al. [23] and the over-classification documented in African American [20–21], US multi-ethnic [19], and Swedish [22] samples and across all five ICMR-NCTB language-specific cutoffs [24]. The emergence and strengthening of the group association at < 24 and < 23 mirror the education-stratified thresholds of Boza-Calvo et al. [32] and the validation by Gupta et al., who identified < 23 as optimal in Indian T2DM (sensitivity 69.2%, specificity 71.8%) [33]. Indian studies have generally applied a single cutoff without comparison (Suvvari et al., < 26, 56% without controls [26]; Gupta et al. 2021, < 23, 22.8% [27]). Part of the steep rise in the prevalence ratio as the cutoff is lowered is the expected behaviour of comparing two separated distributions in their lower tail, and the ratio at < 23 rests on three control events (aPR 6.72; 95% CI 2.23-20.23); we therefore emphasise the absolute excess, whose confidence interval is narrow and far from zero, as the more stable expression of the group difference. This study does not validate < 23 as a diagnostic cutoff, which requires a reference-standard battery; it shows that externally validated thresholds reveal a group association that < 26 obscures, while the glycaemic gradient is visible at all of them.

The diabetic score distribution was both shifted downward and more dispersed than that of controls (variance 10.5 vs. 2.3), and 27.3% of diabetics scored below the lowest control value. Whether this reflects a lower-scoring subgroup within the diabetic group, the relative homogeneity of a comparison group drawn from adults accompanying patients to hospital, or both, cannot be determined from these data; the observation is reported because it explains mechanically why the group contrast is threshold-dependent, not as evidence of a distinct clinical phenotype.

The 44.5% screen-positive yield at < 23 lies close to pooled estimates of mild cognitive impairment in T2DM of 45.0% [34] and 44.1% [35] and within the Indian range [16,36], although those estimates derive from heterogeneous, predominantly elderly (≥ 60 years) populations and from diagnostic rather than screening definitions, so the comparison is indicative only. That a comparable yield appears at a mean age of 50 is consistent with reports of 48.6% in middle-aged South Indian [15] and 59% in 20-60-year-old [14] T2DM patients, and with the excess dementia risk of earlier-onset diabetes [17]. A recent prospective cohort using a gold-standard battery in 832 elderly Indians reported an adjusted OR of 9.1 (95% CI 2.1-39.4) [37]; its comparably large and imprecise estimate mirrors our < 23 result and the rarity of impairment in controls.

The remaining findings are exploratory. The lower abstraction and delayed recall scores, uncorrected across seven subdomains, and the HbA1c correlations concentrated in the same domains, replicate the memory-and-executive pattern of Palta et al. [5] and are biologically plausible given hippocampal vulnerability to insulin resistance, tau hyperphosphorylation, and amyloid-β aggregation [6,9,38] and the frontal white-matter disruption seen on diffusion-tensor imaging in T2DM [39]; the combined amnestic-executive profile carries the highest dementia conversion rate [38]. MoCA subdomains are brief, few-item indices rather than validated domain measures, and no imaging or neuropsychological confirmation was obtained, so these observations are offered as context, not as evidence of a specific pattern of decline. The weak HDL association [43], the absence of a linear LDL association [44], and the absence of a sex difference [35,45] are consistent with the literature but were among many exploratory tests and require replication.

Expressed as triage, < 23 would refer 44.5% of this diabetic clinic and 6.0% of comparable non-diabetic adults for formal neuropsychological evaluation, whereas < 26 would refer over 76% of either group. The first is a discriminating referral fraction; the second is not. What proportion of those referrals would yield confirmed diagnoses is not addressed by these data, because screen-positive status is a trigger for assessment rather than a diagnosis, and this study cannot report the positive predictive value of any threshold. Structured cognitive screening nonetheless remains outside routine care for patients under 65 [13]; whether it should begin earlier in patients with prolonged, poorly controlled T2DM, and at which threshold, is a question these data motivate but cannot settle.

### Strengths and Limitations

Strengths are the continuous, per-unit modelling of HbA1c and duration with a pre-specified test of robustness across three cutoffs, which separates the glycaemic signal from threshold choice; the frequency-matched non-diabetic control group, without which false-positive rates in this population could not be estimated; the restriction to working-age adults with a minimum disease duration, the group that age-based screening guidance currently excludes; and the use of modified Poisson regression, which estimates prevalence ratios directly rather than odds ratios that inflate associations when the outcome is common.

The limitations are substantial. First, no gold-standard neuropsychological battery was administered, so every prevalence figure reported here is a screening yield rather than a disease prevalence, and the data identify a candidate triage threshold rather than validate one. Second, the cross-sectional design precludes causal inference. Third, the control group (n = 50) was modest and imbalanced relative to the diabetic group; the adjusted model at < 23 rests on three control events and its confidence interval (2.23-20.23) is correspondingly wide, so multi-centre replication with larger, balanced samples is needed. Fourth, controls were recruited from adults accompanying patients to the hospital, who may differ from the general population and whose MoCA scores were tightly clustered; the single-centre, hospital-based diabetic sample with a mean HbA1c of 9.25% (78 mmol/mol) represents a poorly controlled clinic population, and community yields may be lower. Fifth, vitamin B12 status was not measured; metformin exposure is near-universal in long-standing T2DM in India, and metformin-associated B12 deficiency is a recognised, reversible contributor to cognitive impairment that could account for part of the duration gradient. Sixth, BMI was calculated from self-reported weight. Seventh, lipid data were unavailable for 24 of 110 diabetics; the near-identical prevalence ratios in the base (aPR 6.72) and lipid-adjusted (aPR 6.81) models suggest minimal bias from the complete-case approach. Eighth, depression was not formally assessed; Gupta et al. found cognitive impairment remained independently associated with cardiometabolic risk factors after adjustment for depression in Indian T2DM [27], and the continuous HbA1c gradient in our data is unlikely to be explained by depressive symptoms alone, but future studies should include a validated depression screen and B12 measurement as covariates.

## CONCLUSIONS

In working-age adults with long-standing, poorly controlled T2DM in a lower-education population, screen-positive cognitive status rose continuously with HbA1c at every MoCA cutoff examined and with diabetes duration at the lower cutoffs, a glycaemic signal independent of threshold choice. The standard cutoff of < 26 classified over three-quarters of both diabetics and controls as screen-positive and could not function as a triage threshold; at < 23, diabetics exceeded matched controls by an absolute 38.5 percentage points (95% CI 25.0 to 48.7). Screen-positive status is not a diagnosis; these data characterise the association and indicate which threshold can triage, not which patients are impaired. Together with independent diagnostic accuracy data [25,33], they support adopting a lower MoCA threshold (< 23) when triaging for cognitive impairment in lower-education Indian T2DM populations, and they make prospective evaluation of earlier structured screening in this group, against a neuropsychological reference standard, a priority.

## Abbreviations

ADA: American Diabetes Association
aPR: adjusted prevalence ratio
BMI: body mass index
CI: confidence interval
HbA1c: glycated haemoglobin
HDL: high-density lipoprotein cholesterol
HR: hazard ratio
ICMR-NCTB: Indian Council of Medical Research Neurocognitive Toolbox
IQR: interquartile range
LDL: low-density lipoprotein cholesterol
MCI: mild cognitive impairment
MoCA: Montreal Cognitive Assessment
NACC: National Alzheimer’s Coordinating Center
OR: odds ratio
PR: prevalence ratio
RR: relative risk
SD: standard deviation
STROBE: Strengthening the Reporting of Observational Studies in Epidemiology
T2DM: type 2 diabetes mellitus

## Acknowledgments

The authors used Claude AI for language editing.

## CRediT author statement

SML: Conceptualization, Methodology, Software, Formal analysis, Investigation, Data curation, Writing-original draft, Visualization. SK: Conceptualization, Methodology, Software, Formal analysis, Investigation, Data curation, Writing-original draft, Visualization. SA: Investigation, Validation, Writing-review & editing. RB: Investigation, Validation, Writing-review & editing. SG: Conceptualization, Validation, Writing-review & editing.

## Data availability

The datasets of the study are available from the corresponding author on reasonable request.

## Ethics approval

This study was approved by the Institutional Ethics Committee of Maulana Azad Medical College, New Delhi and conducted in accordance with the Declaration of Helsinki and its later amendments. Written informed consent was obtained from all participants. Clinical trial number: not applicable

## Consent to Participate

Written informed consent was obtained from all individual participants included in the study.

## Consent for publication

Not applicable; no individual person’s data are reported.

